# Classification of 12-lead ECGs: the PhysioNet/Computing in Cardiology Challenge 2020

**DOI:** 10.1101/2020.08.11.20172601

**Authors:** Erick Andres Perez Alday, Annie Gu, Amit Shah, Chad Robichaux, An-Kwok Ian Wong, Chengyu Liu, Feifei Liu, Ali Bahrami Rad, Andoni Elola, Salman Seyedi, Qiao Li, Ashish Sharma, Gari D. Clifford, Matthew A. Reyna

## Abstract

The subject of the PhysioNet/Computing in Cardiology Challenge 2020 was the identification of cardiac abnormalities in 12-lead electrocardiogram (ECG) recordings. A total of 66,405 recordings were sourced from hospital systems from four distinct countries and annotated with clinical diagnoses, including 43,101 annotated recordings that were posted publicly.

For this Challenge, we asked participants to design working, open-source algorithms for identifying cardiac abnormalities in 12-lead ECG recordings. This Challenge provided several innovations. First, we sourced data from multiple institutions from around the world with different demographics, allowing us to assess the generalizability of the algorithms. Second, we required participants to submit both their trained models and the code for reproducing their trained models from the training data, which aids the generalizability and reproducibility of the algorithms. Third, we proposed a novel evaluation metric that considers different misclassification errors for different cardiac abnormalities, reflecting the clinical reality that some diagnoses have similar outcomes and varying risks.

Over 200 teams submitted 850 algorithms (432 of which successfully ran) during the unofficial and official phases of the Challenge, representing a diversity of approaches from both academia and industry for identifying cardiac abnormalities. The official phase of the Challenge is ongoing.

## Introduction

Cardiovascular disease is the leading cause of death worldwide [1]. Early treatment can prevent serious cardiac events, and the most important tool for screening and diagnosing cardiac electrical abnormalities is the electrocardiogram (ECG) [7, 6]. The ECG is a non-invasive representation of the electrical activity of the heart that is measured using electrodes placed on the torso. The standard 12-lead ECG is widely used to diagnose a variety of cardiac arrhythmias such as atrial fibrillation and other cardiac anatomy abnormalities such as ventricular hypertrophy [7]. ECG abnormalities have also been identified as both short- and long-term mortality risk predictors [9, 4]. Therefore, the early and correct diagnosis of cardiac ECG abnormalities can increase the chances of successful treatments. However, manual interpretation of ECGs is time-consuming and requires skilled personnel with a high degree of training.

The automatic detection and classification of cardiac abnormalities can assist physicians in making diagnoses for a growing number of recorded ECGs. However, there has been limited success in achieving this goal [20, 16] Over the last decade, the rapid development of machine learning techniques have also included a growing number of 12-lead ECG classifiers [21, 15, 3]. Many of these algorithms may identify cardiac abnormalities correctly. However, most of these methods have only been tested or developed in single, small, or relatively homogeneous datasets. In addition, most algorithms focus on identifying a small number of cardiac arrhythmias that do not represent the complexity and difficulty of ECG interpretation.

The PhysioNet/Computing in Cardiology Challenge 2020 provided an opportunity to address these problems by providing data from a wide set of sources with a large set of cardiac abnormalities [5, 12, 13]. The goal of the 2020 Challenge was to identify clinical diagnoses from 12-lead ECG recordings.

We asked participants to design and implement a working, open-source algorithm that can, based only on the clinical data provided, automatically identify any cardiac abnormalities present in a 12-lead ECG recording. We required that each algorithm be reproducible from the provided training data. The winners of the Challenge are the team whose algorithm achieves the highest score for recordings in the hidden test set. We developed a new scoring function that awards partial credit to misdiagnoses that result in similar treatments or outcomes as the true diagnosis or diagnoses as judged by our cardiologists because traditional scoring metrics, such as common area under the curve (AUC) metrics, do not explicitly reflect the clinical reality that some misdiagnoses are more harmful than others and should be scored accordingly.

This year’s Challenge is ongoing. We will update this manuscript with the results after the end of the official phase of the Challenge.

## Challenge Data

For the PhysioNet/Computing in Cardiology Challenge 2020, we assembled multiple databases from across the world. Each database contained recordings with diagnoses and demographic data.

### Challenge Data Sources

We shared data from four sources publicly for training and data from three sources for testing. These sources of ECG data are described below and summarized in Table 1. Two of the three sources of test data are also sources of training data, but few, if any, individuals have ECG recordings in both the training and test sets. We posted the training data and labels, but we did not post the test data or labels. The completely hidden dataset has never been posted publicly, allowing us to assess common machine learning problems such as overfitting.

1. **CPSC**. The first source is the China Physiological Signal Challenge 2018 (CPSC2018), held during the 7th International Conference on Biomedical Engineering and Biotechnology in Nanjing, China [8]. This source includes two databases: a public training dataset (CPSC) and unused data (CPSC-Extra) from CPSC2018. The unused data is not the test data from the CPSC2018, which remains hidden and used for testing in the PhysioNet/CinC Challenge 2020.
2. **INCART**. The second source is the public dataset from the St. Petersburg Institute of Cardiological Technics (INCART) 12-lead Arrhythmia Database, St. Petersburg, Russia, which is posted on in PhysioNet [17].
3. **PTB and PTB-XL**. The third source is the Physikalisch Technische Bundesanstalt (PTB) Database, Brunswick, Germany. This source includes two public databases: the PTB Diagnostic ECG Database [2] and the PTB-XL [18], a large publicly available electrocardiography dataset.
4. **G12EC**. The fourth source is the Georgia 12-lead ECG Challenge (G12EC) Database, Emory University, Atlanta, Georgia, USA. This is a new database, representing a large population from the Southeastern United States, and is split in two for both training and testing.
5. **Undisclosed**. The fifth source is a dataset from an undisclosed American institution that is geographically distinct from the other dataset sources. This dataset has never been (and may never be) posted publicly, and is used for testing in the PhysioNet/CinC Challenge 2020.

**Table 1.** Numbers of patients and recordings in the training databases for the Challenge.

| Source | Patients | Recordings |
| --- | --- | --- |
| CPSC | 9458 | 10330 |
| INCART | 32 | 74 |
| PTB | 19175 | 22353 |
| G12EC | 7871 | 10344 |

### Challenge Data Variables

Each annotated ECG recording contained ECG signal data and demographic information, including age, sex, and a diagnosis or diagnoses, i.e., the labels for the Challenge data.

Table 2 provides a summary of the age, sex, and recording information for the training data, indicating differences between the populations. Table 3 and Figure 1 provide summaries of the diagnoses for the training data. The training data contain 111 diagnoses/classes, where we used 27 of the 111 total diagnoses to evaluate participant algorithms. These 27 diagnoses were relatively common, of clinical interest, and more likely to be recognizable from ECG recordings. The list of scored diagnoses for the Challenge can be seen in Table 3 with long-form descriptions, corresponding SNOMED CT codes, and abbreviations. Only these scored classes are shown in Table 3 and Figure 1, but all 111 classes are included in the training data so that participants can decide whether or not to use them with their algorithms. The test data contain a subset of the 111 diagnoses in potentially different proportions, but each diagnosis in the test data is represented in the training data.

**Table 2.**
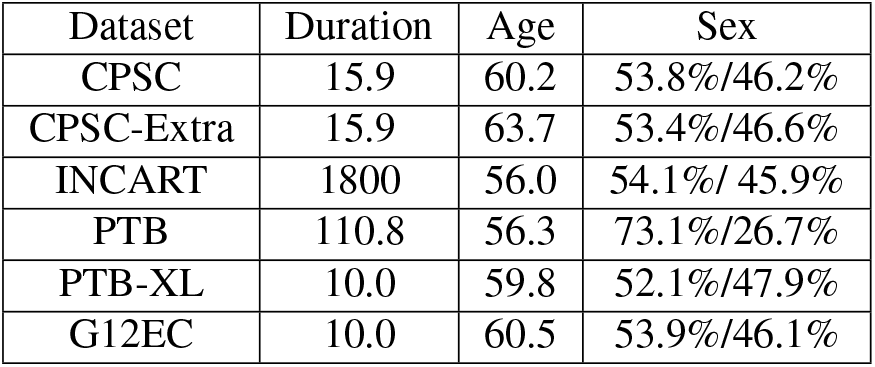
Mean duration of recordings in years, mean age of patients in recordings in seconds, and sex (male/female) of patients in recordings for each training set.

| Dataset | Duration | Age | Sex |
| --- | --- | --- | --- |
| CPSC | 15.9 | 60.2 | 53.8%/46.2% |
| CPSC-Extra | 15.9 | 63.7 | 53.4%/46.6% |
| INCART | 1800 | 56.0 | 54.1%/ 45.9% |
| PTB | 110.8 | 56.3 | 73.1%/26.7% |
| PTB-XL | 10.0 | 59.8 | 52.1%/47.9% |
| G12EC | 10.0 | 60.5 | 53.9%/46.1% |

**Table 3.**
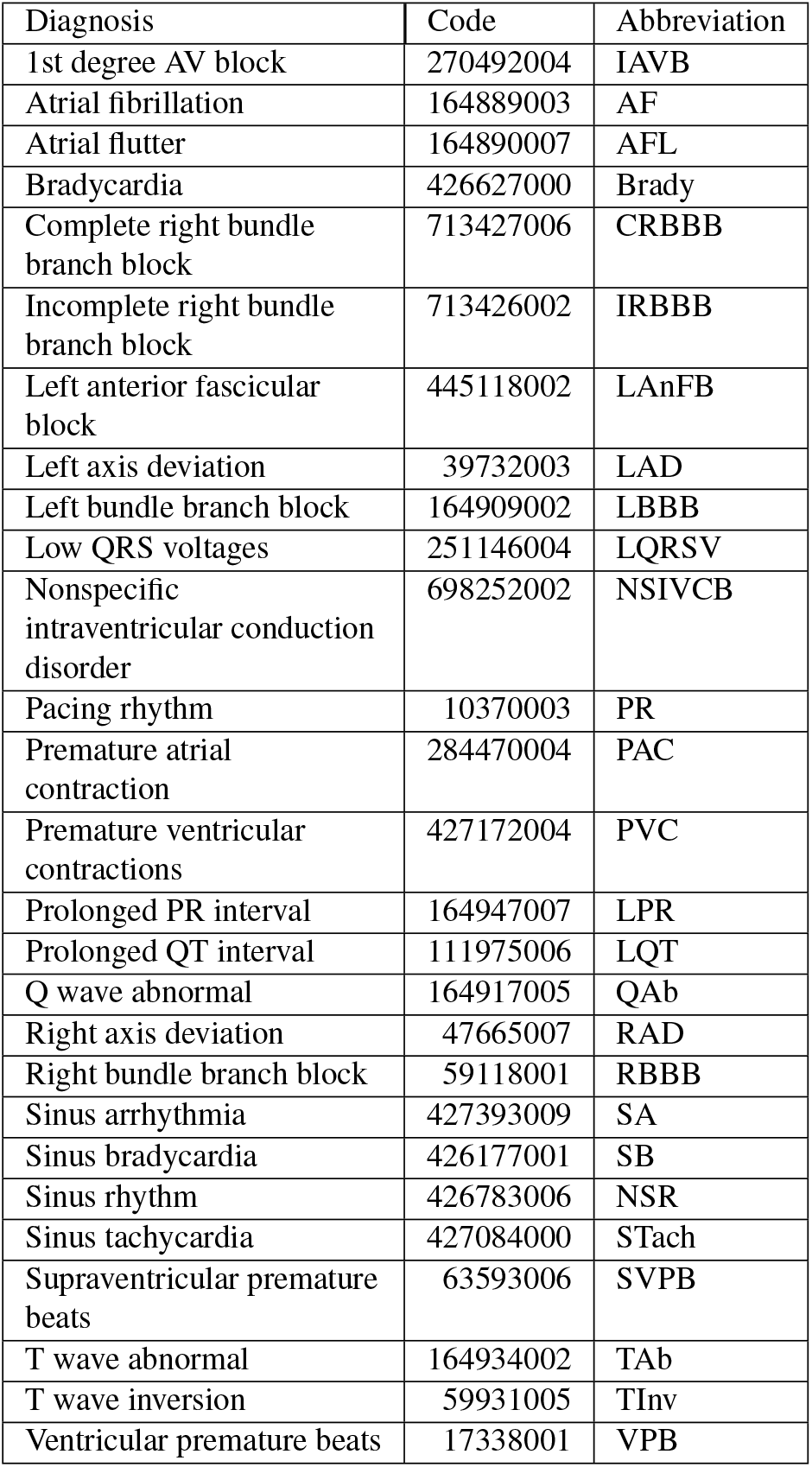
Diagnoses, SNOMED CT codes and abbreviations in the posted training databases for diagnoses that were scored for the Challenge.

**Fig. 1.**
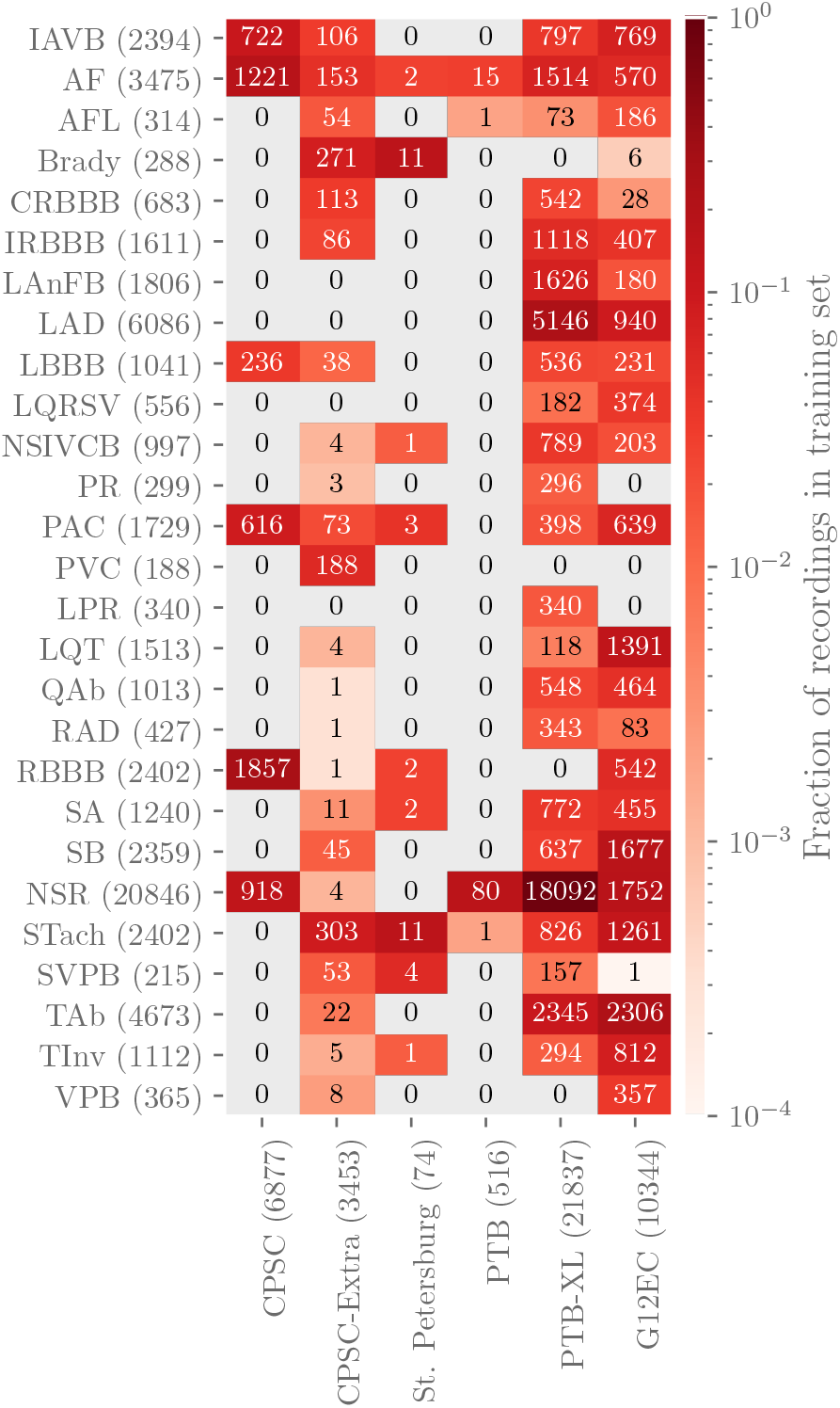
Numbers of recordings with each scored diagnosis in each training set with colors indicating the fraction of recordings with each scored diagnosis in each training set, i.e., the total number of each scored diagnosis in a training set normalized by the number of recordings in the training set. Parentheses indicate the total numbers of records with a given label across training sets (rows) and the total numbers of recordings, including recordings without scored diagnoses, in each training set (columns).

All data were provided in WFDB format [5]. Each ECG recording had a binary MATLAB v4 file for the ECG signal data and a text file in WFDB header format describing the recording and patient attributes, including the diagnosis or diagnoses, i.e., the labels for the recording. We did not change the original data or labels from the databases, except (1) to provide consistent and Health Insurance Portability and Accountability Act (HIPPA)-compliant identifiers for age and sex, (2) to add approximate SNOMED CT codes as diagnoses for each recording, and (3) to change the amplitude resolution to save the data as integers as required for WFDB format.

## Challenge Objective

We asked participants to design working, open-source algorithms for identifying cardiac abnormalities in 12-lead ECG recordings. To the best of our knowledge, for the first time in any public competition, we required code both for a team’s trained models and the code training their models to aid the generalizability and reproducibility of the research conducted during the Challenge. We ran the participants’ trained models on the hidden test data and evaluated their performance using a novel, expert-based evaluation metric that we designed for this year’s Challenge.

### Challenge Overview, Rules, and Expectations

This year’s Challenge is the 21^st^ PhysioNet/Computing in Cardiology Challenge [5]. Like previous Challenges, this year’s Challenge had an unofficial phase and an official phase. The unofficial phase (February 7, 2020 to April 30, 2020) provided an opportunity to socialize the Challenge and sought discussion and feedback from teams about the data, evaluation metrics, requirements. The unofficial phase allowed 5 scored entries for each time. After a short break, the official phase (May 11, 2020 to August 23, 2020) introduced additional training and test data, a requirement for teams to submit their training code, and an improved evaluation metric. The official phase allowed 10 scored entries for each team. During both phases, teams were evaluated on only a small subset of the test data; evaluation on the full test set occurs after the end of the official phase of the Challenge to prevent sequential training on the test data. Moreover, while teams are encouraged to ask questions, pose concerns, and discuss the Challenge in a public forum, they are prohibited from discussing their particular approaches to preserve the uniqueness of their approaches to solving the problem posed by the Challenge.

### Classification of 12-lead ECGs

We required teams to submit both their trained models along with code for training their models. (We announced this requirement at the launch of this year’s Challenge but did not start requiring the submission of training code until the official phase of the Challenge.) Teams included any processed and relabeled training data in this step; any changes to the training data are part of training a model.

We first ran each team’s training code on the full training data and then ran each team’s trained code from the previous step on sequentially on recordings from the hidden test sets.

We allowed teams to submit either Python or MATLAB implementations of their code. Other languages, including R and Julia, were supported but received insufficient interest from participants during the unofficial phase. Participants containerized their code in Docker and submitted it using GitHub or Gitlab. We downloaded their code and ran in containerized environments on Google Cloud. The computational environment is described more fully in [14] from the previous year’s Challenge.

We used virtual machines on Google Cloud with 8 vCPUs, 64 GB RAM, and an optional NVIDIA T4 Tensor Core GPU with a 72 hour time limit for training on the full training set. We used virtual machines on Google Cloud with 2 vCPUs, 13 GB RAM, and an optional NVIDIA T4 Tensor Core GPU with a 24 hour time limit for running the trained classifiers on the full test set.

To aid teams, we shared baseline models that we implemented in Python and MATLAB. The Python baseline model was a random forest classifier that used age, sex, QRS amplitude, and RR intervals as features. QRS detection was implemented using the Pan-Tompkins algorithm [10]. The MATLAB baseline model was a hierarchical multinomial logistic regression classifier that used age, sex, and global electrical heterogeneity [19] parameters as features. The global electrical heterogeneity parameters were computed using a time coherent median beat and origin point calculation [11].

### Evaluation of Classifiers

For this year’s Challenge, we developed a new scoring metric that awards partial credit to misdiagnoses that result in similar outcomes or treatments as the true diagnoses as judged by our cardiologists. This scoring metric reflects the clinical reality that some misdiagnoses are more harmful than others and should be scored accordingly. Moreover, it reflects the fact that it is less harmful to confuse some classes than others because the responses may be similar or the same.

Let 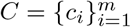 be a collection of *m* distinct diagnoses for a database of *n* recordings. First, we defined a multi-class confusion matrix *A =* [*a_ij_*], where *a_ij_* is the normalized number of recordings in a database that were classified as belonging to class *ci* but actually belong to class *c_j_* (where *c_i_* and *c_j_* may be the same class or different classes). Since each recording can have multiple labels and each classifier can produce multiple outputs for a recording, we normalized the contribution of each recording to the scoring metric by dividing by the number of classes with a positive label and/or classifier output. Specifically, for each recording *k* = 1,…,*n*, let *x_k_* be the set of positive labels and *y_k_* be the set of positive classifier outputs for recording *k*. We defined a multiclass confusion matrix *A =* [*a_ij_*] by

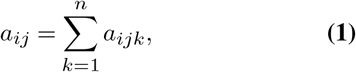

where

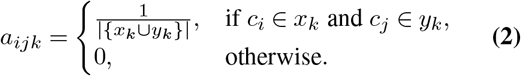

The quantity |{*x_k_* ⋃ *y_k_* }| is the number of distinct classes with a positive label and/or classifier output for recording *k*. To incentivize teams to develop multi-class classifiers, we allowed classifiers to receive slightly more credit from recordings with multiple labels than from those with a single label, but each additional positive label or classifier output may reduce the available credit for that recording.

Next, we defined a reward matrix *W =* [*w_ij_*], where *w_ij_* is the reward for a positive classifier output for class *ci* with a positive label *c_j_* (where *c_i_* and *c_j_* may be the same class or different classes). The entries *W* are defined by our cardiologists based on the similarity of treatments or differences in risks (see Table 4). The highest values of the reward matrix are along its diagonal, associating full credit with correct classifier outputs, partial credit with incorrect classifier outputs, and no credit for labels and classifier outputs that are not captured in the weight matrix. Also, three similar classes (i.e., PAC and SVPB, PVC and VPB, CRBBB and RBBB) are scored as if they were the same class, so a positive label or classifier output in one of these classes is considered to be a positive label or classifier output for all of them. However, we did not change the labels in the training or test data to make these classes identical to preserve any institutional preferences or other information in the data.

**Table 4.**
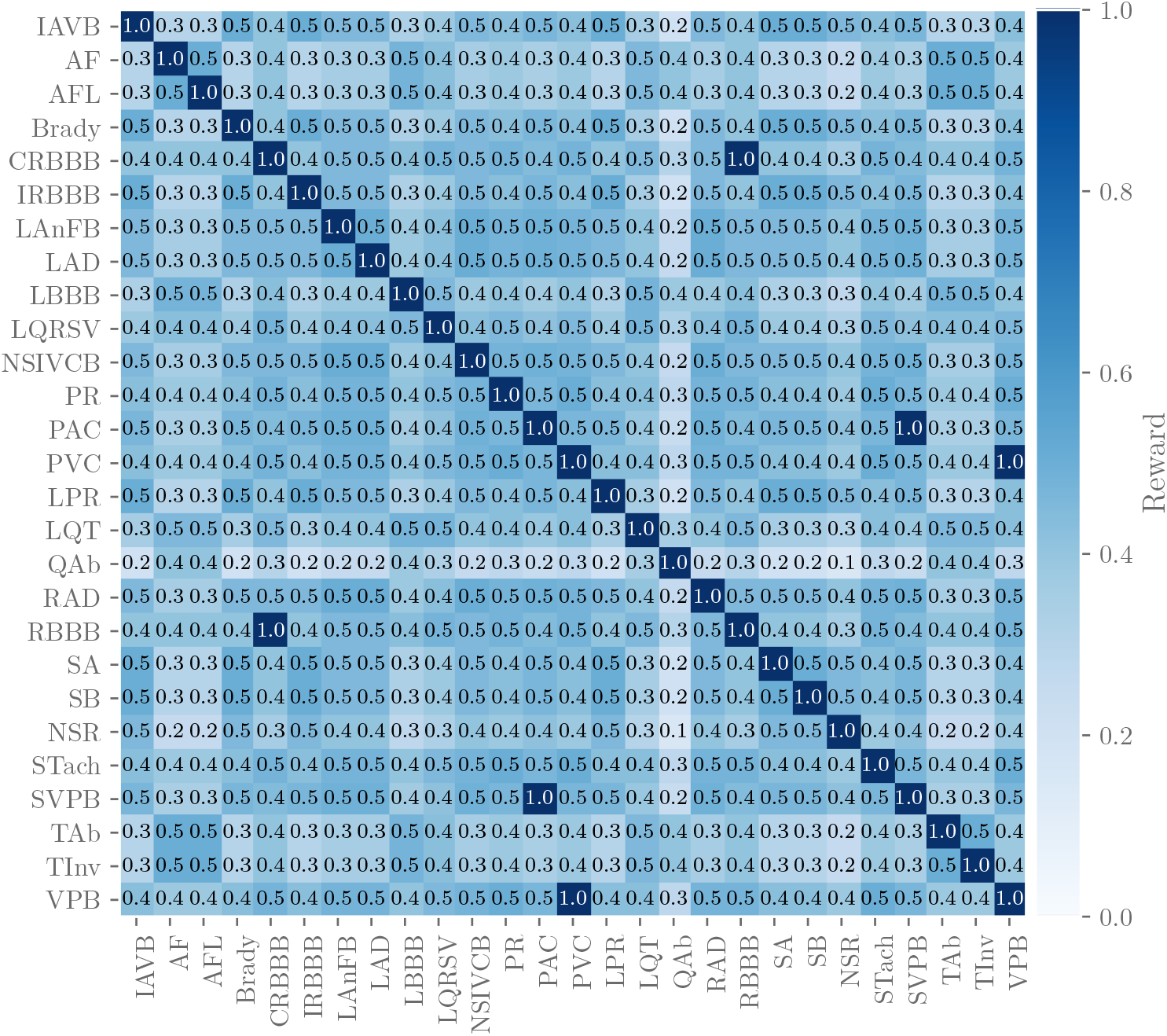
Reward matrix *W* for the diagnoses scored in the Challenge with rows and columns labeled by the abbreviations for the diagnoses in Table 3. Off-diagonal entries that are equal to 1 indicate similar diagnoses that are scored as if they were the same diagnosis. Each entry in the table was rounded to the first decimal place due to space constraints in this manuscript, but the shading of each entry reflects the actual value of the entry.

Finally, we defined a score

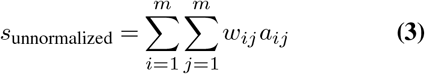

for each classifier as a weighted sum of the entries in the confusion matrix. This score is a generalized version of the traditional accuracy metric that awards full credit to correct outputs and no credit to incorrect outputs. To aid interpretability, we normalized this score so that a classifier that always outputs the true class or classes receives a score of 1 and an inactive classifier that always outputs the normal class receives a score of 0, i.e.,

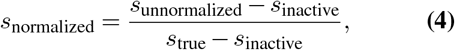

where *s*_inactive_ is the score for the inactive classifier and *s*_true_ is the score for ground-truth classifier. A classifier that returns only positive outputs will typically receive a negative score, i.e., a lower score than a classifier that returns only negative outputs, which reflects the harm of false alarms.

Accordingly, this scoring metric is designed to award full credit to correct diagnoses and partial credit to misdiagnoses with similar risks or outcomes as the true diagnosis. The resources, populations, practices, and preferences of an institution all determine the ideal choice of the reward matrix *W*; the choice of *W* for the Challenge is just one example.

## Discussion

At the time of writing, we have received over 850 submissions of algorithms from over 200 teams across academia and industry. The most common algorithmic approach was based on deep learning and convolutional neural networks. However, over 70% of entries used standard clinical or hand-crafted features with classifiers such as support vector machines, gradient boosting, random forests, and shallow neural networks.

## Conclusions

This article describes the world’s largest open access database of 12-lead ECGs. The data were drawn from three continents with diverse and distinctly different populations, encompassing 111 diagnoses. We have also introduced a novel scoring matrix that rewards algorithms based on similarities between diagnostic outcomes, weighted by severity/risk.

The public training data and sequestered test data provided the opportunity for unbiased and comparable repeatable research. To the best of our knowledge, this is the first public competition that has required the teams to provide both their original source code and the framework for (re)training their code. In doing so, this creates the first truly repeatable and generalizable body of work on classification of electrocardiograms.

## Data Availability

All posted training datasets are available through the Challenge website at https://physionetchallenges.github.io/2020/.

## Acknowledgements

This research is supported by the National Institute of General Medical Sciences (NIGMS) and the National Institute of Biomedical Imaging and Bioengineering (NIBIB) under NIH grant number 2R01GM104987-09 and R01EB030362 respectively, the National Center for Advancing Translational Sciences of the National Institutes of Health under Award Number UL1TR002378, as well as the Gordon and Betty Moore Foundation and AliveCor, Inc. under unrestricted gifts. Google also donated cloud compute credits for Challenge teams. GC has financial interest in Alivecor and Mindchild Medical. GC also holds a board position in Mindchild Medical. Neither of these entities influenced the design of the Challenge or provided data for the Challenge. AIW holds equity and management roles in Ataia Medical and is supported by the NIGMS 2T32GM095442. AE receives financial support from the Basque Government through grant PRE_2019_2_0100. The content of this manuscript is solely the responsibility of the authors and does not necessarily represent the official views of the above entities.

